# Displaying emergency patient estimated wait times: a multi-centre, qualitative study of patient, community, paramedic and health administrator perspectives

**DOI:** 10.1101/2020.08.11.20173153

**Authors:** Katie Walker, Melanie Stephenson, Jennie Hutton, Anne Loupis, Keith Joe, Michael Ben-Meir, Ella Martini, Michael Stephenson, Judy Lowthian, Beatrice Yip, Elena Wu, James Ho, Gabriel Blecher, Kim Hansen, Paul Buntine

## Abstract

**Background:** Emergency Departments have the potential ability to predict patient wait times and to display this to patients and other stakeholders. Little is known about whether consumers and stakeholders would want this information and how wait time predictions might be used. The aim of this study was to gain perspectives from consumer, referrer and health services personnel regarding the concept of emergency wait time visibility.

**Methods:** In 2019, 103 semi-structured interviews and one focus group were conducted with emergency medicine patients/families, paramedics, well community members and hospital/paramedic administrators. Nine emergency departments and multiple organisations in Victoria, Australia, contributed data. Transcripts were coded and themes are presented.

**Results:** Consumers and paramedics face physical and psychological difficulties when wait times aren’t visible. Consumers believe about a 2-hour wait is tolerable, beyond this most begin to consider alternative strategies for seeking care. Consumers want to see triage to doctor times; paramedics want door to off-stretcher times (for all possible transport destinations); with 47/50 consumers and 30/31 paramedics potentially using this information. Twenty-eight of 50 consumers would use times to inform facility or provider choice, 19/50 want information once in the waiting room. During prolonged waits, 1/52 consumers would consider not seeking care. Visibility of approximate waits would better inform decision-making, improve load-spreading, allow planning and access to basic needs and might reduce anxiety.

**Conclusions:** Consumers and paramedics want wait time information visibility. They would use the information in a variety of ways, both pre-hospital and whilst waiting for care.

## Introduction

The decision to seek emergency medical care is complex. Many factors influence what care to seek and when. Once a facility or provider is chosen, acutely unwell or distressed patients hope to see a doctor/definitive clinician immediately on arrival. This is rarely achievable; most patients join a queue. Previously pre-hospital providers and patients have been blinded to queue duration. Emergency Department (ED) proximity and wait times have a major influence on patient choice of facility(1-3). Patient satisfaction is lower during longer waits and also when the actual wait is longer than the expected wait(4, 5). The reverse is also true.

In the era of expanding information technology (IT) capabilities, there is much time-based data available to hospitals. People are increasingly IT literate and some seek mobile information to inform decision-making. In the USA, wait estimates are widely available and in Australia, there are individual centres and jurisdictions making real-time ED wait time information public. In efforts to predict wait-times, algorithms have been developed and tested for accuracy(6-8). This isn’t straightforward and some algorithms are inaccurate(9, 10). Researchers have begun work on system-wide load-spreading for patients once ED capacity is predicted(11).

Little is known about what patients, pre-hospital and hospital workers feel about wait time visibility nor about the impact of wait time transparency. A single-centre survey demonstrated that American patients strongly supported wait time visibility(12). An opinion regarding how patients might interpret predicted wait times has been published without any evidence(13). Health personnel worry that giving people information might place them at risk of making decisions not to seek care because queues are long(14). A before and after study looked at the impact of wait time visibility for two hospitals, finding lengthy waits reduced with visible wait times(3). Before health administrators implement visible wait times in our communities, more information is needed on consumer and health care worker perspectives.

The aim of this study was to gain perspectives from consumer, referrer and health services personnel regarding emergency wait time visibility. This included exploring patient safety, defining wait times important to consumers and paramedics and determining whether and how people would use predicted wait times if they were available.

## Methods

### Study design

This qualitative interview and focus group study was conducted between July and November 2019 to determine consumer, community, pre-hospital and hospital administrator perspectives on emergency department wait time visibility. We employed a purposeful sampling strategy and used a grounded theory approach, using semi-structured interviews. The study was undertaken in Melbourne, Australia.

The study received Cabrini ethics committee approval (10-13-05-19) and was prospectively registered on ANZCTR.org.au: ACTRN12619000665134.

### Characteristics of interviewer

The female interviewer was an emergency trainee introduced as a university researcher, whilst wearing office attire, with no previous relationship with participants.

### Settings and participant selection

We sought information from six groups: patients/support persons in EDs, paramedics, well community members, paramedic controllers, hospital administrators and general practitioners (GPs). EDs were invited to participate if they were part of an academic health science centre (regional to tertiary hospitals). Other hospitals were recruited via researcher networks. Recruitment methodology for each participant group is described in Figure 1. Paramedics and controllers were recruited from Ambulance Victoria. GPs were invited to participate in phone interviews during education sessions for GPs at Cabrini. Community members included (i) sporting club members, (ii) special/preschool/primary/secondary school parents and (iii) older people receiving nursing home services from a large district home nursing services provider. Face-to-face recruitment was by convenience sampling.

**Figure 1.**
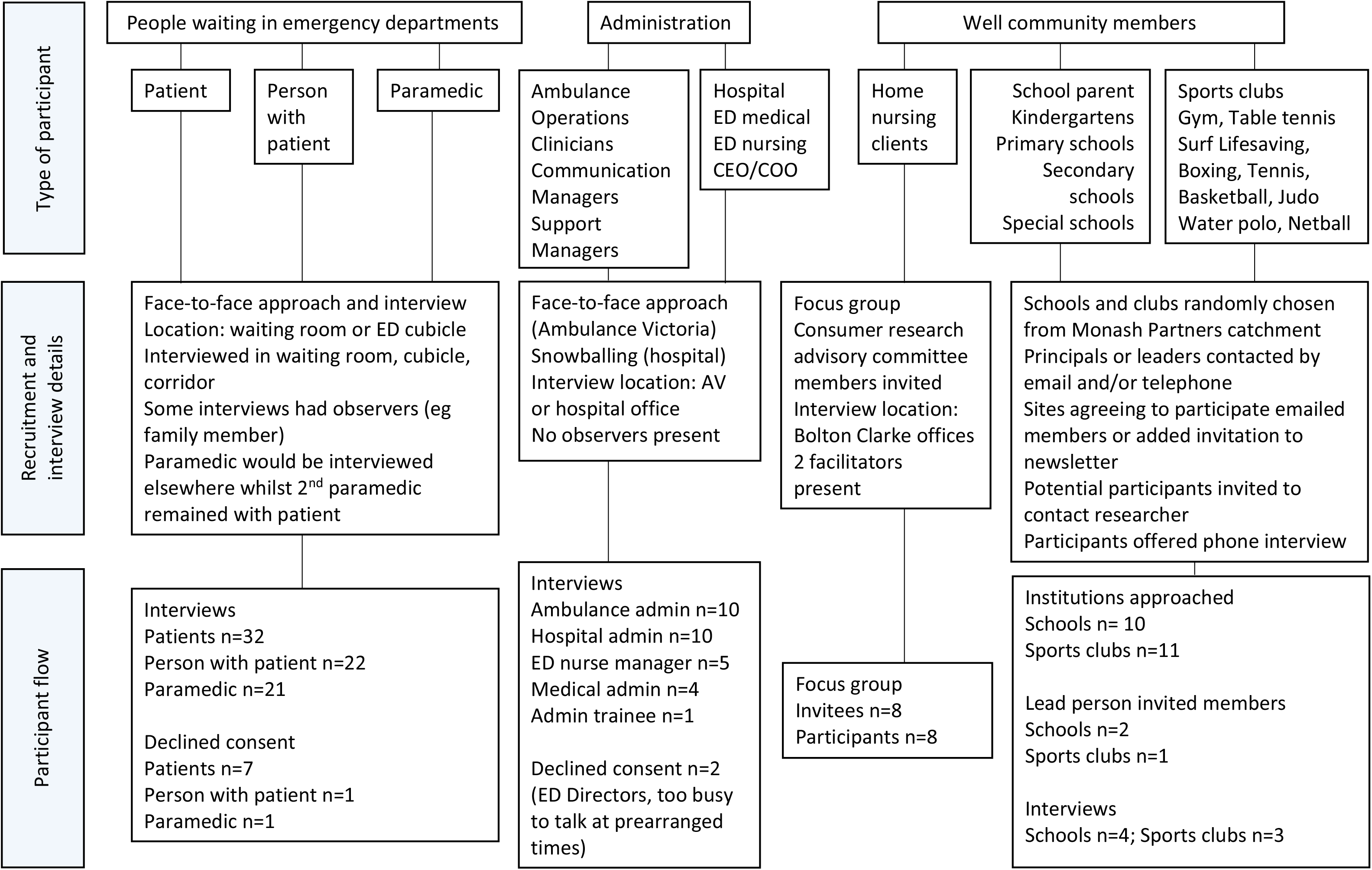
Recruitment methodology and flow through the study

Sampling continued until thematic and information saturation was achieved, then three further interviews were obtained from each group.

### Data collection

Interviews and the focus group were semi-structured with potential questions and prompts as needed (Appendix 1). There were no repeat interviews or transcription checking by participants. Interviews were recorded as audio files for professional transcription. Field notes were undertaken during the focus group. Interview duration ranged from 15 to 30 minutes.

### Data analysis and findings

Four researchers coded interviews using NVivo software (QSR, Melbourne, Australia) or paper. They worked independently, coding transcripts using open coding techniques. An iterative process was undertaken, refining the codes until most content could be classified. Themes were developed using an inductive and iterative approach and content and agreement on themes was reached at a researcher meeting.

## Results

### Flow through the study

One-hundred and three interviews were conducted and eight people participated in a single home nursing service focus group. Eight people in EDs declined interviews. Few community members responded to invitations and no GPs participated. One transcript was lost (demographics unknown). All transcribed interviews were included in the study. (Figure 1.)

### Hospitals and individual participants

Participating EDs included two medium metropolitan, three large metropolitan, two major, one major paediatric and one private hospital. (Table 1.) One hospital displays a wait time online, to Ambulance Victoria control, and in its ED waiting room (Cabrini). Four hospitals declined to participate (major, medium and large metropolitan, private). Interview participants are described in Table 1. Interviewees had a median age of 49 (range 20,93, IQR 33,58) and 59 (55%) were female.

**Table 1.** Demographics of participating interviewees and emergency departments

| Table 1. Demographics of interview participants |  |  |  |
| --- | --- | --- | --- |
| Type of participant | Participants (n) | Age (median, IQR) | Female (n, %) |
| Patient in ED | 32 | 51 (31,70) | 20, 65% |
| Carer or advocate in ED | 22 | 52 (44,58) | 17, 77% |
| Paramedic in ED | 21 | 31 (27,44)† | 12, 60%† |
| Ambulance Victoria controllers | 10 | 54 (49,55) | 2, 20% |
| Hospital Administrator | 10 | 52 (41,58) | 5, 50% |
| Sports club | 3 | mean=50, (N/A) | 1, 33% |
| School parent | 4 | 44 (39,46) | 2, 50% |
| General Practitioner | 0 | N/A | N/A |
| Home nursing service client | 8 | 76 (68, 83) | 4, 50% |
| **Total** | **111** | **49 (33,58)** | **59, 55%** |
| †Demographics not available for one paramedic |  |  |  |
| Emergency Department/Type* | Participants (n)<br>patients/advocates | Attendances per annum |  |
| Angliss, medium metropolitan | 5 | 41,000 |  |
| Box Hill, large metropolitan | 10 | 67,000 |  |
| Cabrini, private | 13 | 23,000 |  |
| Casey, medium metropolitan | 4 | 65,000 |  |
| Clayton, major (paediatrics and adult) | 6 | 70,000 |  |
| Dandenong, large metropolitan | 6 | 87,000 |  |
| Maroondah, large metropolitan | 5 | 60,000 |  |
| St Vincent's, major | 5 | 46,000 |  |
| **Total** | **54** | **459,000** |  |

\*Australian Institute of Health and Welfare descriptor

### Themes

Thematic saturation was achieved for patients, their advocates/carers and for ambulance administrators and paramedics. Theme saturation wasn’t achieved for hospital administrators or community members (parents of school children, sports club members, home nursing service clients). Themes relating to ED wait time visibility are presented below. Themes related to predicted ED wait time visualisation are presented in another manuscript.

### Perspectives on not having a visible ED wait time

Consumers in the ED wanted to know their estimated wait time 48/51 (94%). Three consumers (6%) were happy to wait for care regardless of wait duration and didn’t wish to know how long this might be.

Consumers report feeling a lack of control when the wait time isn’t visible. They arrive at an ED seeking immediate treatment and then face a void of information following triage. Patients reported wondering if they should go home, feeling that if they had a true emergency, they would have been treated immediately. Some find their anxiety increases the longer they have to wait and discussed how having a wait time visible in the ED waiting room would lower their anxiety and assist them with planning other commitments.

*“People get cross and angry, three hours, that’s a long time to be ignored” (consumer)*

Others worry about planning their next few hours, whilst having competing demands on their time.

*“I’ve got three kids…(it’s)annoying sitting there and thinking do I need to get someone to pick them up?” (consumer)*

Many consumers commented that if wait times were visible, they could manage their wait better (including toileting, food, family/work communications, psychological expectations).

*“I could have got a coffee or gone to the bathroom” (consumer)*

Consumers report that after an hour of waiting, they start asking the triage nurse if they are still in the queue.

Health administrators consider that consumers have the right to transparent information about wait times and many hoped to make times available.

*“I think if publicising wait times helps patients make choices” (health administrator)*

Patients similarly requested wait time transparency. Paramedics remarked that transparency would reduce patient anxiety whilst waiting. Paramedics also commented that knowing wait times would help with planning care whilst patients remained on stretchers (eg toileting needs, administering pain relief, changing oxygen bottles, paramedic shift changes). (Table 2.)

**Table 2.** Perspectives on not having a visible wait time prediction

|  |
| --- |
| ***Information Void*** |
| You wouldn't know if you had time to get something (to eat or drink) because you don't know how long you have to wait.<br>*Consumer 25M I66* |
| I think transparency and clarity about what's going on helps people. I think people often feel that they just don't know what's going on. They've had one point of contact they speak to someone and then they're just sitting there. *Health Administrator 30M I62* |
| It's mentally draining when you're in the room and don't know what's going on. That's pretty much what it feels like the nurses don't know, the whole hospital doesn't know. *Paramedic 27F I75* |
| ***Transparency*** |
| I think the more people feel as if they are receiving information that's relevant and timely it creates that trust, it will grow.<br>*Consumer 51M I25* |
| You feel like you're really shut out and have no information, you don't know how to escalate. *Consumer 47F I2* |
| I think communication and education is really important for people in the queue. People get cross and angry, three hours that's a long time to feel ignored. You lose faith in the department then in the hospital. I think you just get disappointed that no one cares about your child. You don't know if everyone has gone on their coffee break. *Community 47F I2* |
| Important to be visible and to be transparent so that the resources are allocated in the right spots. *AV 52M I5* |
| I think if publicising wait times helps patients make choices and it's reasonably accurate you have to start somewhere.<br>*Hospital Administrator 55M I17* |
| Paint an actual factual picture I would prefer them to say it's Armageddon here. But they don't. They go around and paint a pretty picture potentially that just doesn't deliver. *AV 60M I11* |
| ***Anxiety management*** |
| The communication of wait times makes such a difference. Makes people calm down and it's okay at least you're recognising we have been here for three hours. *Consumer 58F I64* |
| It would ease their calm a little bit that they know they've got a time. *Paramedic 29M I23* |
| **Planning** |
| With little ones at home just even managing the expectation of whoever is at home with them how long. *Consumer 49F I14* |
| I've got three kids so I would need to know how long I'm going to be gone for. There's nothing more annoying than sitting there and thinking do I need to get someone to pick them up, get a babysitter, do this or that. *Consumer 33F I29* |
| We were in a rush today so I only bought one bottle. We're here for two feeds I had to find a kitchen boiling water to rinse a bottle and fill it again. It's important to plan. *Consumer 33F I29* |
| If I want to go out to the car make a private phone call and I know I can't be seen (for a while) then I can use my time. You daren't move from the waiting room in case they call your name and you slip back (in the queue). I'm sure everyone would say that. *Consumer 62F I28* |
| I could've got a coffee or bathroom. *Consumer 26F I63* |
| (I would like the) wait time to help plan. I know I've got a one-hour car park here. *Consumer 49M I84* |
| I would be able to get some food (for the patient). It's about making sure the patient is cared for. *Paramedic 29M I34* |
| You don't know how long you're going to be have to sustain (a patient) on the stretcher. If they need to go to the toilet or things like that. *Paramedic 58F I86* |
| **Asking the triage nurse about progress in the queue** |
| I mean after an hour and a half I will go back up and see whether they've forgotten about us. *Consumer 54M I33* |
| You have to go and ask have to go and say where am I at. Then you feel bad because you know they're busy but you go to the counter and say how long do you think. That's not fun for them to get that all night from different people as well. *Community 47F I2* |
| I did ask at one stage because I needed to know how to manage my patient, if they were going to go straight to a bed, I wouldn't need to give more pain relief. We often need to know to organise another crew to come and relieve us. When I asked, I had my head bitten off quite nastily. I didn't feel very comfortable in approaching again. I just wanted a bit of communication. *Paramedic 48F I24* |

*I = Interview number*

*“I did ask at one stage because I needed to know how to manage my patient, if they were going to go straight to a bed, I wouldn’t need to give more pain relief. We often need to know to organise another crew to come and relieve us.” (paramedic)*

### Expectations about waiting in an ED

Thirty-one consumers voiced opinions about how long it was reasonable to wait to see a provider. Seven thought less than one hour, 13 thought 1-2 hours and 11 were comfortable waiting longer than 2 hours. Most consumers interviewed had already been waiting longer than two hours.

*“I would think one hour would be a very long waiting time, realistically, it can be two hours” (consumer)*

Once waits were longer than 2 hours, consumers either felt they would wait as long as it took or were considering an alternative to their local ED. Alternatives considered were primary care or representing at a quieter time. Consumers who had been waiting over 6 hours seemed accepting that there was a prolonged wait and felt safer sitting in the ED waiting room than at home or in their GPs. (Table 3.)

**Table 3.** Expectations about waiting in an ED

|  |
| --- |
| ***Willingness to wait*** |
| I guess I'm prepared to wait until I'm seen. *Consumer 44F I73* |
| If it was something that could wait until I could see my GP I wouldn't be in here. I need to be seen. *Consumer 42F I78* |
| No longer. We've done five hours here, we're done. *Consumer 27F I81* |
| ***Seeking alternatives to the ED waiting room*** |
| I might go to another place. Seek an alternative. *Consumer 51M I25* |
| Four hours I probably would just go home and if it got bad I would call an ambulance. *Consumer 40F I92* |
| You could say I could go home ring 13 SICK (GP after hours service) and that might be a better solution for tonight. *Community 47F I2* |
| Give them their options. Tell them how long the wait is in a hospital but make sure they're aware they're welcome to come to that hospital. *Hospital Administration 61F I62* |
| ***Expectations*** |
| I think it (*wait time visibility*) can have a negative impact because sometimes it can set expectations. *Hospital Administration 61F I62* |
| From a patient's point of view, the positive would be that they're given an upfront ballpark figure as to how long they might expect to wait. A patient's expectation may not be able to match what is realistic. *Hospital Administrator 58M I110* |

*I = Interview number*

### Impact and decision making if ED wait times are visible

All consumers in ED waiting rooms perceived that they had an urgent condition needing treatment at an ED. If prolonged wait times were made visible, consumers discussed: choosing an ED with a shorter wait time; presenting to their closest ED anyway as they felt they would be treated quickly (queues were for everyone else); considering alternatives to ED treatment; calling an ambulance (anticipating a shorter queue) or attending their usual hospital, regardless of wait, due to a long-term relationship with the facility.

*“I would travel further to not wait as long, definitely” (consumer)*

*“if it was crazy wait times and your patient was low-acuity, you could (go) to another hospital” (paramedic)*

*“it would just be the closest hospital” (consumer)*

Paramedics reported being more interested in which was the closest appropriate facility than in the length of the wait. They said that if their protocols required it, they would be happy to choose the facility with the shortest wait time provided the hospital could meet the patient’s needs and the consumer accepted the decision.

Health administrators felt that consumers had capacity to integrate wait time information into appropriate decision making and that it would assist them in choosing a provider.

*“it’s paternalistic of the profession not to trust patients to make valid decisions on where they want to go” (administrator)*

One administrator felt that all patients should stay in their local area to facilitate post-hospital integrated care and family support and hence wait times weren’t relevant to their population. (Table 4.)

**Table 4.** Potential impact of wait time visibility

|  |
| --- |
| ***Shortest wait time*** |
| I would travel further to not wait as long, definitely. *Consumer 31F I21* |
| It depends on the situation. If it's life threatening or critical you're not going to care what the wait time is you just need to be seen. If it's less concerning but just knowing, you can bring something with you to entertain yourself or if it's a relatively short wait time. *Consumer 26F I63* |
| If I could compare hospitals in my area and say this one has a six hour wait time this is a two hour wait time, I would probably go to the two hour wait time. *Community 47F I2* |
| If it was crazy wait times and your patient was low acuity it could be useful because then you could reload to a different hospital. *Paramedic 34M I34* |
| Often, we find ourselves with a patient that doesn't have any history at a particular hospital but we're sort of in between two hospitals in terms of distance, time to ED. *Paramedic 31F I70* |
| By being able to give someone the information of look (local) Hospital is a five and a half hour wait your local GP is a great source of knowledge and appropriate for today, it's more appropriate to see them. *Paramedic 27F I87* |
| ***Closest hospital*** |
| For a full emergency it would just be the closest. *Community 35M I1* |
| For me it's more about where is the closest or the most appropriate to go for the care. We actually came in last night and I didn't even think to look anywhere else. I just knew that here was accessible and we've had good experience here before. *Consumer 31F I21* |
| ***Call an ambulance*** |
| Go see 24-hour Doctor first see if we can get a referral to come straight in or if it's really an emergency ring an ambulance and come through with an ambulance. There are the two options rather than having to wait four or five hours. *Consumer 52M I64* |
| It's how long will I have to wait if I go in my car and often that's their motivation in calling an ambulance because they think they can jump the queue. *AV 54F I8* |
| **No change** |
| There's probably a large cohort of patients that have history with a certain hospital where you think it's probably best they stay at that hospital. *Paramedic 33F I16* |
| I think on road we would still be expected to go to the closest ED. *AV 54F I8* |
| **Decision making** |
| If I'm going to be admitted, I'd rather be where they've got my history. But If I've something urgent I will go to the closest hospital. *Community 41 FG4* |
| I can just foresee a little bit of perhaps angst in here and a lot more conversations over the air in here about people perhaps arguing or wanting to go to different hospitals. *AV 54F I8* |
| I think it's somewhat paternalistic and arrogant of the profession not to trust patients to make valid decisions on where they want to go. *Hospital Administration 48M I13* |
| I feel like it's sort of good information that it does empower patients to make decisions. *Hospital Administration 31F I37* |

*I = Interview number*

### Safety issues regarding visibility

One out of 52 consumers stated that they would go home and not seek further care if wait times were many hours. The rest stated that they would wait for care, return the next day or seek alternate care (another ED, GP or other provider).

*“if I thought something was seriously wrong, I’d stay however long it took” (consumer)*

*“(long waits) wouldn’t stop me from going to see somebody but it might change where I go” (consumer)*

Most consumers understand that patients with time critical illness don’t wait and wait times don’t apply in this scenario. There was a variety of knowledge presented by consumers regarding triage and different waits for different people. Health administrators and paramedics expressed concerns about long wait predictions potentially deterring patient from seeking help. (Table 5.)

**Table 5.** Safety issues regarding wait time visibility

|  |
| --- |
| **Safety** |
| It might make me re-evaluate my need to go. It wouldn't stop me from going to see somebody but it might change where I go. *Community 35M I1* |
| I would be seen quicker in emergency if I felt I needed to go to emergency I would definitely come. If I knew that I'm going to wait four hours then I probably wouldn't come. *Consumer 58M I64* |
| I think I would make a personal assessment of my condition. If I felt like it was critical condition or heart related or something really wasn't right it would influence where I go. *Consumer 31F I21* |
| You feel safe when you're in the hospital. We're prepared to wait. *Consumer 52F I19* |
| If I thought something was seriously wrong, I'd stay however long it took. *Consumer 27F I71* |
| I'm here because I need to be seen by a clinician. If it was something that could wait until I could see my GP I wouldn't be in here. *Consumer 42F I78* |
| if I've got to come I've got to come. *Consumer 71M I91* |
| The only thing I can see being adverse is if they self-triage based on a hospital ED wait time. I think if anybody is truly critically unwell then they shouldn't be triaging themselves because that's not how acuity works, you get seen sooner. *Paramedic 44M I69* |
| I probably disagree with it because I think there would be some patients who would not go to hospital if they perhaps need to because they don't understand the triage process. *Paramedic I85* |
| So I think it's an important part of information that a consumer needs and needs to be accurate. You could artificially direct people away particularly if they're long wait times. I think people should be able to go to their closest provider but make an intelligent choice. People might be driving dangerously across town with chest pain to go somewhere that's publicised as five minutes as opposed to half an hour. *Hospital Administration 55M I17* |
| Say I was bleeding had a heart attack or if the kids were sick my immediate reaction wouldn't be let's see where is the quietest but more where is the closest hospital and how do I get there. *Consumer 35M I1* |
| **Health Literacy regarding ED queues and wait times** |
| If you come in and I have at times come in with breathing problems or pains in the chest or something as such you sort of go straight in. *Consumer 71M I91* |
| If my face is split open, I have no worries waiting for someone who has a life-threatening condition basically. *Consumer 43M I18* |
| It's not fair for me because I logged in first 45 minutes and someone that actually needs more emergency level and she logged in half an hour after me and has to wait over one hour I don't think that's fair. *Consumer 37M I82* |
| I don't know how you prioritise. *Consumer 62F I28* |
| You get triaged on how priority it is *Consumer 33F I29* |
| Wait times applies to anybody who presents at emergency that needs to see a Doctor. *Consumer 27F I71* |

*I = Interview number*

### Definition of an ED wait time?

One hundred and one participants expressed opinions on this. The majority of non-ambulance participants wanted triage to doctor times displayed. Consumer interviewees defined a start time as when triage commenced (33/52) or when they arrived at the front door (18/52). One thought the time started when they got to the car park. The finish time was when consumers saw their doctor/definitive clinician (30/51). The majority of hospital administrators and community members felt the same (triage to doctor). A quarter of consumers wanted the total ED time to be provided (triage to discharge) (11/52). Very few people from any group wanted to know when they would go to a cubicle or have management commenced (medical triage, nursing assessment, medications, imaging, pathology). (See Appendix 2. for ED journey stages)

Ambulance personnel wanted the waiting time to start when they arrived at the door (22/31), with (9/31) feeling the start time was the triage time. Most (30/31) felt the finish time was when the patient was off-loaded from their stretcher (ie door to off-stretcher duration).

### Would people use visible estimated ED wait times?

Of those asked, 47/50 of consumers felt they would use a wait time estimator. Twenty-eight of fifty consumers said they would look up a wait time prior to attending an ED as this would inform their choice of facility. Another 19/50 said they would look at the wait time on arrival as they needed to get to an ED before doing anything else (including researching where to go). Almost all wanted the wait time to be available in waiting rooms. Three interviewees didn’t want to see wait times at any stage of their journey. Well community members supported wait time visibility (6/7) as did hospital administrators (8/9).

Thirty of thirty-one ambulance employees (paramedics and controllers) felt they would use a wait time estimator. Twenty-five would use times to assist with load-spreading (choosing hospitals with shorter waits), three would use the time once in an ED, one wouldn’t use the information. Importantly, paramedics will only use wait time predictions for load-spreading if all hospitals in the region are supplying information and they are available in one place electronically for comparison.

## Discussion

This study shows the potential value of ED wait time displays to multiple stakeholders. Patients can better manage anxiety associated with their wait; physical needs such as toileting and food; family enquiries and external responsibilities such as work and family obligations. Wait times could assist ambulance services with managing distribution and will assist with caring for patients in stretcher queues within EDs. Health service administrators could use wait times to manage consumer expectations and to monitor performance. Almost all stakeholders would use this information, either prior to attending an ED, in the waiting room, or at both times.

Consumers felt around 2 hours was a reasonable wait time. If prolonged predicted wait times were displayed, consumers would have varied strategies for obtaining care (including seeking alternate sources of care) or they would stay and wait in their ED of choice but could plan to use their time better with less anxiety. Health administrators felt patients would be able to use wait time information to better inform decisions. Consumers advised about long waits almost always considered an alternate provider, rather than not seeking care. Patients wanted to see triage to main clinician times (25% preferred triage to discharge), ambulance personnel wanted door to off-stretcher times. 99% of consumers and 97% of paramedics would use a wait time predictor at some time during the patient journey.

To our knowledge, this is the first study seeking broad input from consumers and ED stakeholders on the concept of displaying predicted wait times. The study largely contradicts a theoretical emergency medicine paper raising risk concerns about the use of wait time predictors(13) and agrees with Shaikh’s findings from Baltimore(12). The 1/52 consumers that felt that prolonged waits would make them consider going home and not seeking care is consistent lower than the “did-not-wait” rates at most EDs(15, 16).

The study supports the importance of managing wait times to optimise patient satisfaction. In a systematic review of contributors to ED patient satisfaction, wait time reduction (actual and perceived) was the second most important theme (of fifteen)(17). Two studies have shown that perceived wait time (not actual wait time) is the most important variable contributing to patient satisfaction(4, 18). Patients and their families don’t have very accurate perceptions of the duration of their waits(19). Waits of uncertain length feel longer than waits of certain lengths(20), providing estimated wait times could shorten the perceived wait.

The study was multicentre, incorporating information from many types of ED and interviewing a diverse group of stakeholders. The study should be generalisable regarding consumers and ambulance employees within Australia and may or may not be more widely generalisable.

### Limitations

Participants were from a single city in Australia. The data were from a mixture of interviews and focus groups, findings from one method of data collection may differ from another. The study didn’t achieve theme saturation for health administrators, well community members or GPs, so there may be views that haven’t been presented in this jurisdiction, or different views in other locations. In particular, health administrators may have barriers to transparency that haven’t been mentioned (such as concerns about highlighting long waits or receiving more patients due to shorter waits). Future work should consider the views of emergency healthcare workers. The study doesn’t report on the best visualisation of data or how to safely display data. A comprehensive safety evaluation would require post-implementation studies. Wait time predictions would depend on EDs collecting and analysing electronic metadata in real-time (not currently universally available). Interviews presumed that predicted wait times are accurate, how consumers respond to inaccurate predictions hasn’t been explored.

### Conclusion

Consumers and paramedics want wait time information visibility. They would use the information in a variety of ways, both pre-hospital and whilst waiting for care. Wait time visibility is likely to improve the patient experience of emergency medicine.

## Data Availability

The data is not available to the public due to confidentiality concerns of the participants.

## Acknowledgements

Lisa Kuhn, Cathie Piggott, Anne Spence - governance assistance.

## Collaborators

Rachel Rosler (Monash Health)

Hamish Rodda (Eastern Health)

David Rankin (Cabrini Health)

John Papatheohari (Cabrini Health)

## Competing interests

Some authors are emergency physicians or directors, others work in community health (pre-hospital and district nursing). One author is a consumer. KW is a section editor for EMA.

## Funding

The Medical Research Future Fund, via Monash Partners, funded this study. Researchers contributed in-kind donations of time. The Cabrini Institute provided research infrastructure support and the Cabrini Foundation provided the salary for Mel S.

## Contributions

Funding: KW, MBM, KJ

Study protocol: KW, Mel S, Mick S, JL, JH, EM, KH

Site CIs: GB, Mel S, PB, JH, JL

Ethics: Mel S, KW

Data collection: Mel S, BY, EW

Data analysis: AL, BY, EW, Mel S, KW

Manuscript draft: KW, AL, BY, EW

Revisions: All authors

Manuscript guaranteed by KW

## Appendix 1. Interview guide

### Questions for Patient/Accompanying persons in Emergency Department

#### Wait times actual vs expectation

When do you think the ED wait time should begin?

When do you think the ED wait time ends?

What do you think about waiting times to see an emergency doctor?

Prompts:

How long have you been waiting in this ED today?

What wait time were you hoping for when you arrived?

Were you given any information on your arrival about the approximate wait time today?

Do you know how long it will be from now until you see the doctor?

Would you have liked any information about your likely wait time?

#### Advertised wait times – awareness/impact

What do you think about being able to see how long the wait time is for emergency departments?

When did you decide to attend this ED?

Did you know that Internet wait time information or waiting room wait time information exists for some EDs?

Do you have access to the Internet via a computer or mobile phone?

Did you look for wait-time information before choosing this ED?

Would this be important to you next time you are unwell?

Would you like to see this before you get to the ED? Would you like this information when you arrive at the ED?

#### Type/Accuracy information

What do you think about the accuracy of wait time information?

Prompts:

How important is accuracy of the wait time to you?

What impact would waiting longer than the advertised wait time have on you?

What impact would waiting less than the advertised wait times have on you?

How would you like times presented? (use actual examples from the web in Victoria)

What sort of approximation of the wait time prediction would be acceptable to you?

Prompts:

Accurate to within 5 minutes/30 minutes/1 hour or >1 hour?

Would you like to be given the time to seeing a doctor or time to seeing a nurse or both?

Would you like to have other information available?

Prompts:

Advice that critically unwell patients get seen immediately, despite the advertised wait

Wait times may change whilst you are waiting

Other places that care can be sought for various conditions

That a triage nurse will start your treatment when you arrive

#### Stratification/understanding of severity?

Who do you think advertised ED wait times apply to?

Would wait times influence your decision regarding which facility to attend?

Prompt: If there is a long wait, will this alter your decision-making? How?

Please expand?

How long are you prepared to wait to be seen by a clinician today?

Prompt:

5 mins/15 mins/30 mins/1 hour/2 hours or >2 hours?

Would you consider leaving without being seen? When? Why or why not?

Would you seek alternate medical help? Why or why not?

Any other comments?

### Questions for Paramedics

#### Wait times actual vs expectation

When do you think the ED wait time should begin?

When do you think the ED wait time ends?

How long have you been waiting in this ED today?

Were you given any information on your arrival about the approximate patient wait time today?

Would you have liked any information about the patient’s likely wait time?

Ambulance offload time? Or both?

#### Advertised wait times – awareness/impact

Did you know that Internet wait time information or waiting room wait time information exists for some EDs?

Do you have access to the Internet (on your phone) when choosing an emergency department?

Did you look for wait-time information before choosing this ED?

Would you like to see this before you get to the ED? Would you like this information when you arrive at the ED?

How important is accuracy of the ED wait time to you?

What impact would waiting longer than the advertised wait time have on you?

What impact would waiting less than the advertised wait times have on you?

How would you like times presented? (use actual examples from the web in Victoria)

What sort of approximation of the wait time prediction would be acceptable to you eg accurate to within 5 minutes/30 minutes/1 hour or >1 hour?

What sort of wait time information would you like to see? Time to offload/time to see a nurse/time to see a doctor or something else?

#### Decision making

Would ED wait times influence your decision regarding which facility to attend?

If there is a long ED wait, will this alter your decision-making? How? Please expand? Any further comments?

**For AV controllers/Diversion Officer only:**

What do you think the barriers and enablers are to distribute patients to departments with capacity?

Questions for Health administrators:

1. When do you think the ED wait time should begin?
2. When do you think the ED wait time ends?
3. What do you think the positive and negative impacts of advertised Emergency Department wait times are on:

a. Patients
b. Ambulance services
c. Emergency departments
d. Individual Hospitals
e. State wide health service

Prompts:

Health Transparency

Patient Safety

Shared decision making

Any perceived barriers

4. What sort of approximation of the wait time prediction would be acceptable to you eg accurate to within 5 minutes/30 minutes/1 hour or >1 hour?
5. Would you like to have other additional information available?
6. How do you feel this information is best presented?
7. Would you like to advertise the time to seeing a doctor, time to seeing a nurse or something else?
8. What potential issues would you have with your service publishing wait times to the following groups:

- Ambulance Victoria
- Emergency Department waiting rooms
- Internet
9. Any further comments?

### Questions for Community members

#### Advertised wait times – awareness/impact

Did you know that Internet wait time information or waiting room wait time information exists for some EDs?

When do you think the ED wait time should begin?

When do you think the ED wait time ends?

Do you have access to the Internet via a computer or mobile phone?

Did you look for wait-time information before choosing your last attendance at an Emergency Department?

Would this be important to you next time you are unwell?

Would you like to see this before you get to the ED?

Would you like this information when you arrive at the ED?

How important is accuracy of the wait time to you?

What impact would waiting longer than the advertised wait time have on you?

What impact would waiting less than the advertised wait times have on you?

How would you like times presented? (use actual examples from the web in Victoria)

What sort of approximation of the wait time prediction would be acceptable to you eg accurate to within 5 minutes/30 minutes/1 hour or >1 hour?

Would you like to be given the time to seeing a doctor or time to seeing a nurse or both?

Would you like to have other information available?

Prompt if required, potential discussion points below:

- Advice that critically unwell patients get seen immediately, despite the advertised wait
- Wait times may change whilst you are waiting
- Other places that care can be sought for various conditions
- That a triage nurse will start your treatment when you arrive

#### Stratification/understanding of severity?

Who do you think advertised ED wait times apply to?

Would wait times influence your decision regarding which facility to attend?

If there is a long wait, will this alter your decision-making? How? Please expand. Any other comments?

---

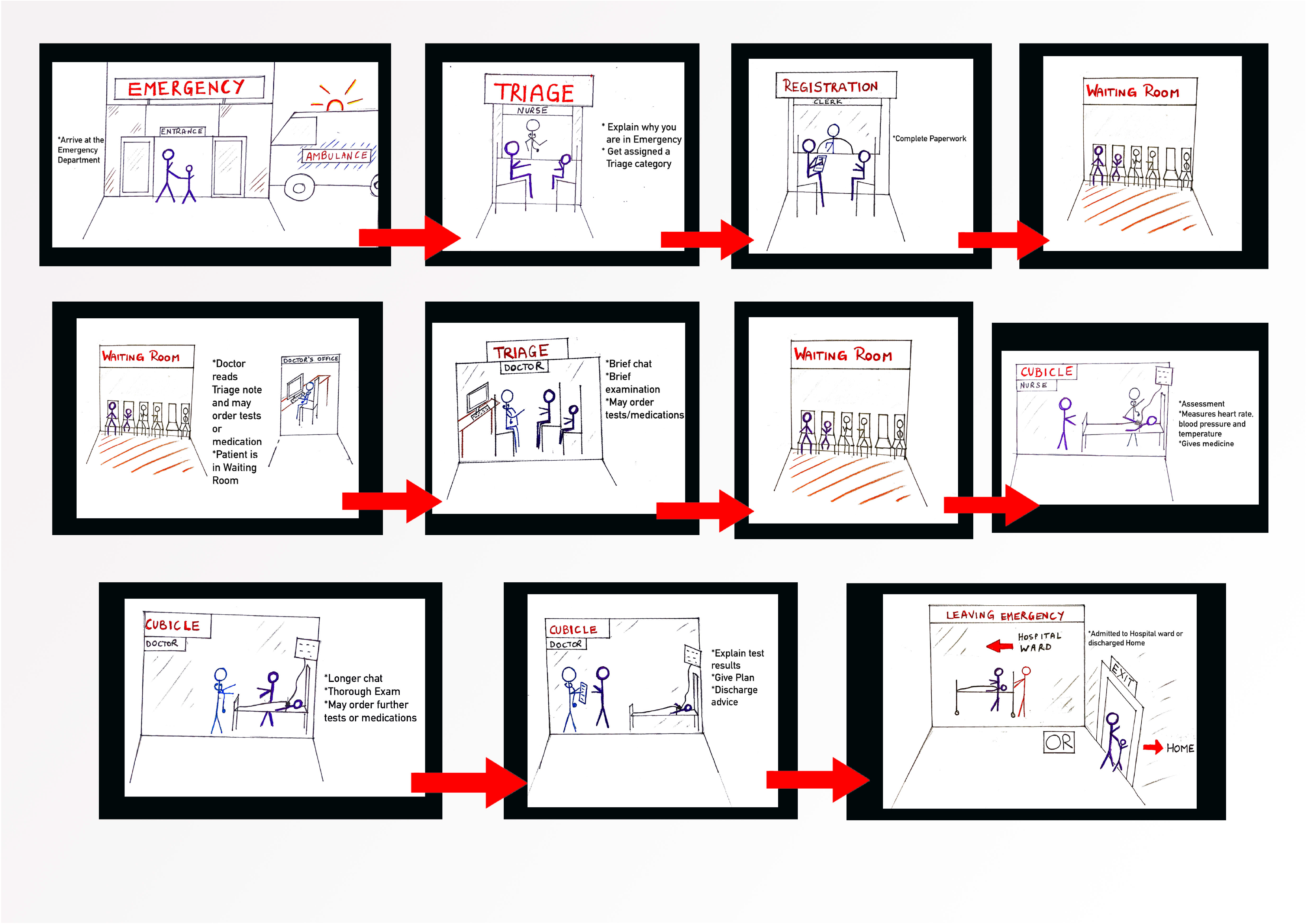

## Notes

### Clinical Trial

ACTRN12619000665134

### Author Declarations

The study received Cabrini ethics committee approval (10-13-05-19)

